# Nowcasting food insecurity on a global scale

**DOI:** 10.1101/2021.06.23.21259419

**Authors:** Giulia Martini, Alberto Bracci, Sejal Jaiswal, Matteo Corea, Lorenzo Riches, Jonathan Rivers, Elisa Omodei

**Affiliations:** World Food Programme, Research, Assessment and Monitoring Division (RAM), Via Cesare Giulio Viola 68, 00148 Rome, Italy; Department of Mathematics, City, University of London, EC1V 0HB, London, UK

## Abstract

Lack of regular physical or economic access to safe, nutritious and sufficient food is a critical issue affecting millions of people world-wide. Estimating how many and where these people are is of fundamental importance for governments and humanitarian organizations to take informed and timely decisions on relevant policies and programmes. In this study, we propose a machine learning approach to predict the prevalence of people with insufficient food consumption and of people using crisis or above crisis food-based coping when primary data is not available. Making use of a unique global data set, we show that the proposed models can explain up to 78% of the variation in insufficient food consumption and crisis or above food-based coping levels. We also show that the proposed models can be used to nowcast the food security situation in near-real time and propose a method to identify which variables are driving the changes observed in predicted trends, which is key to make predictions serviceable to decision makers.

---

Characterizing the socioeconomic status of populations and providing reliable and up-to-date estimates of who the most vulnerable are, how many they are, where they live and why they are vulnerable is essential for governments and humanitarian organizations to make informed and timely decisions on policies and programmes [1]. This data is traditionally collected through face-to-face surveys. However, these are expensive, time-consuming, and in certain areas not possible to perform due to conflict, insecurity or remoteness. Therefore, during the last few years, researchers have begun to investigate the potential of non-traditional data and new computational methods to estimate vulnerabilities and socioeconomic characteristics when primary data is not available. In these studies, mobile phone data [2], satellite imagery [3], a combination of both [4, 5], geolocated Wikipedia articles [6] or Tweets [7], and social media advertising data [8], have been used in combination with state-of-the-art machine learning methods to provide reliable estimates of poverty at different spatial resolutions for several Sub-Saharan African countries as well as Southern and Southeastern Asian ones.

The methods proposed in these studies provide a unique opportunity to monitor poverty in near real-time on a global scale. In this work, we show that similar methods and data can be used to tackle another outstanding form of vulnerability affecting populations world-wide: food insecurity. In 2019, the number of under-nourished people was estimated to be almost 690 millions [9], with 135 millions in 55 countries and territories reported to be acutely food insecure [10]. These numbers have significantly increased as a consequence of the COVID-19 pandemic, with at least 155 million people reported to be acutely food insecure in 2020, an increase of around 20 millions from the previous year [11]. In order to address this global issue, monitoring the situation and its evolution is key. Governments and international organizations like the World Food Programme (WFP), the Food and Agriculture Organization (FAO) and the World Bank perform food security assessments on a regular basis through face-to-face surveys or, increasingly so, remote mobile phone surveys (e.g. computer-assisted telephone interviews, CATI) and further supporting technologies such as interactive voice response and web surveys [1]. However, as mentioned previously, there are limitations with these approaches given their high costs in both monetary and human resources. In addition, food insecurity is a more dynamic and unstable phenomenon than poverty, with a seasonal component related to agricultural production calendars, and subject to swift changes when external shocks hit, therefore requiring more frequent and rapid assessments.

Food insecurity is a multidimensional concept, spanning from food availability and access to utilization and stability [12]. Multiple indicators have been developed to characterize household food insecurity levels, each capturing different aspects. In this study we focus on the Food Consumption Score (FCS) and the reduced Coping Strategy Index (rCSI), the former capturing quantity and diversity of dietary intake and the latter the consequences of constrained access to food, resulting in coping behaviors [13]. Aggregating a representative number of household-level measurements of these indicators makes it possible to characterize the food security situation of a geographical area during a specified time window through the prevalence of people with insufficient food consumption and that of people using crisis or above crisis food-based coping, respectively. In this study, we show that these two metrics can be estimated from secondary data by means of machine learning algorithms, when primary data is not available. This opens the door to food security near-real time nowcasting on a global scale, allowing decision makers to make more timely and informed decisions on policies and programmes oriented towards the fight against hunger.

Previous work has explored the use of secondary data to investigate specific aspects of food insecurity, such as agricultural production. Statistical crop models and climate modeling have been used to make projections for 2030 of changes in crop productions in 12 food-insecure regions due to climate change [14]. Mobile phone records have been used to analyze monthly mobility in Senegal leading to the discovery and characterization of seasonal mobility profiles related to economic activities, agricultural calendars and precipitation [15]. Other studies have proposed a characterization of the food security situation based on a variety of secondary indicators, addressing its multidimensional aspect and providing annual national-level estimates [16, 17]. Famine risk prediction through machine learning and stochastic models has also been the subject of recent investigation. Okori and Obua used household socioeconomic and agricultural production characteristics to train several machine learning models to predict households’ food security status [18]. The limitation of this approach is that up-to-date household level data is required not only during model training but also when using the trained models to perform out-of-sample predictions. More recently, the World Bank developed a suite of statistical models to forecast transitions into critical states of food insecurity and famine risk from secondary data [19, 20]. In this study, we focus on food security nowcasting, proposing a methodology that allows, for the first time, to estimate the current prevalence of people with insufficient food consumption and of people using crisis or above crisis food-based coping at sub-national level at any given time from secondary data, when primary data is not available. Seminal work by Lentz and collaborators addressed this challenge for the first time, obtaining satisfactory predictions for food consumption, although limited to Malawi only [21]. Here, we make use of a unique data set of sub-national level food consumption and food-based coping data collected during the last 15 years across, respectively, 78 and 41 countries (see Figure 1), allowing for the first time the development and validation of nowcasting predictive models of food security indicators on a global scale.

**Figure 1:**
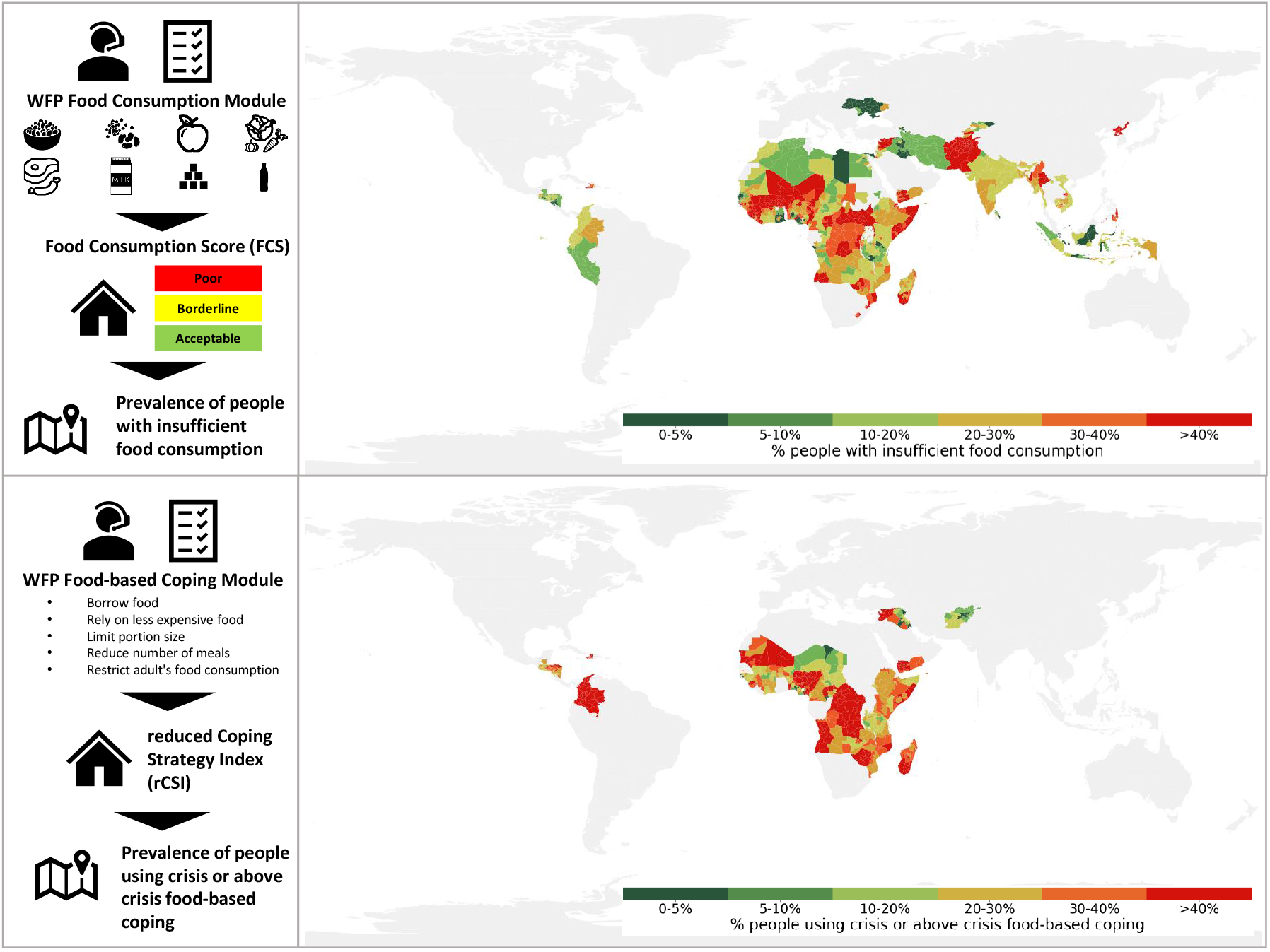
Geographical distribution of insufficient food consumption and crisis of above crisis food-based coping data. The two left panels summarize the data collection and aggregation process: operators collect household-level food consumption and coping strategies information through face-to-face or mobile phone surveys, from which the Food Consumption Score (FCS) and the reduced Coping Strategy Index (rCSI) are calculated for each household. By aggregating this information across a representative number of surveys, the prevalence of people with insufficient food consumption and the prevalence of people using crisis or above crisis food-based coping in a given geographical area and time window is obtained. The two right panels show the geographical distribution of the sub-national (first-level administrative units) assessments collected by WFP, governments and other organizations during the last 15 years in 78 and 41 countries, for food consumption and food-based coping respectively. Each area is colored by the severity of the latest measured prevalence as defined by WFP [22].

## Results

### Predicting levels of insufficient food consumption and of crisis or above food-based coping from previously measured levels and secondary data

The main assumption of this study is that, when primary data is not available, levels of insufficient food consumption and of crisis or above food-based coping can be estimated from secondary information, specifically on the key drivers of food insecurity. Experts identify three main causes for food insecurity: conflict, economic shocks, and extreme weather events [10]. To build the proposed predictive models, we therefore collected historical data covering all three dimensions: data on conflict-related fatalities, economic information (prices of staple food in local markets, headline and food inflation, currency exchange rates, and GDP per capita) and data on rainfall and vegetation, including anomalies with respect to historical averages. For each available historical measurement of insufficient food consumption and of crisis or above food-based coping for a given geographical area and time window, we associate as independent variables the corresponding conflict, economic and weather situation for the same area in the previous three-month window. Moreover, we also take into account as independent variables undernourishment and population density, as well as the target prevalence measured during the previous food security assessment.

For each target variable, we fitted *N*_*b*_ = 100 bootstrapped logistic regression models using gradient boosted regression trees [23], employing a random 80% sample of the historical data, as further detailed in the *Methods*. As reported in Table 1, the proposed models are able to explain, on the remaining 20% out-of-sample data, 75% of the variation in the prevalence of people with insufficient food consumption and 78% of the variation in the prevalence of people using crisis or above crisis food-based coping, with a mean absolute error of, respectively, 0.08 and 0.06. Figure 2 (top plots) shows the predicted vs actual prevalence for each observation in the test set. The former is calculated as the median of the predicted values obtained from the *N*_*b*_ bootstrapped models.

**Table 1:** Model performance metrics. Coefficients of determination (*R*^2^) and mean absolute errors (MAE) obtained on the test set for each of the four proposed models and for the naive approach, which simply uses the prevalence measured during the previous assessment as the predicted prevalence.

| | | $R^2$ | MAE |
| --- | --- | --- | --- |
| Food consumption | Prevalence from previous assessment included as independent variable | 0.75 | 0.08 |
|  | Prevalence from previous assessment **not** included as independent variable | 0.63 | 0.09 |
|  | Naive model | 0.39 | 0.12 |
| Food-based coping | Prevalence from previous assessment included as independent variable | 0.78 | 0.06 |
|  | Prevalence from previous assessment **not** included as independent variable | 0.73 | 0.07 |
|  | Naive model | 0.42 | 0.10 |

**Figure 2:**
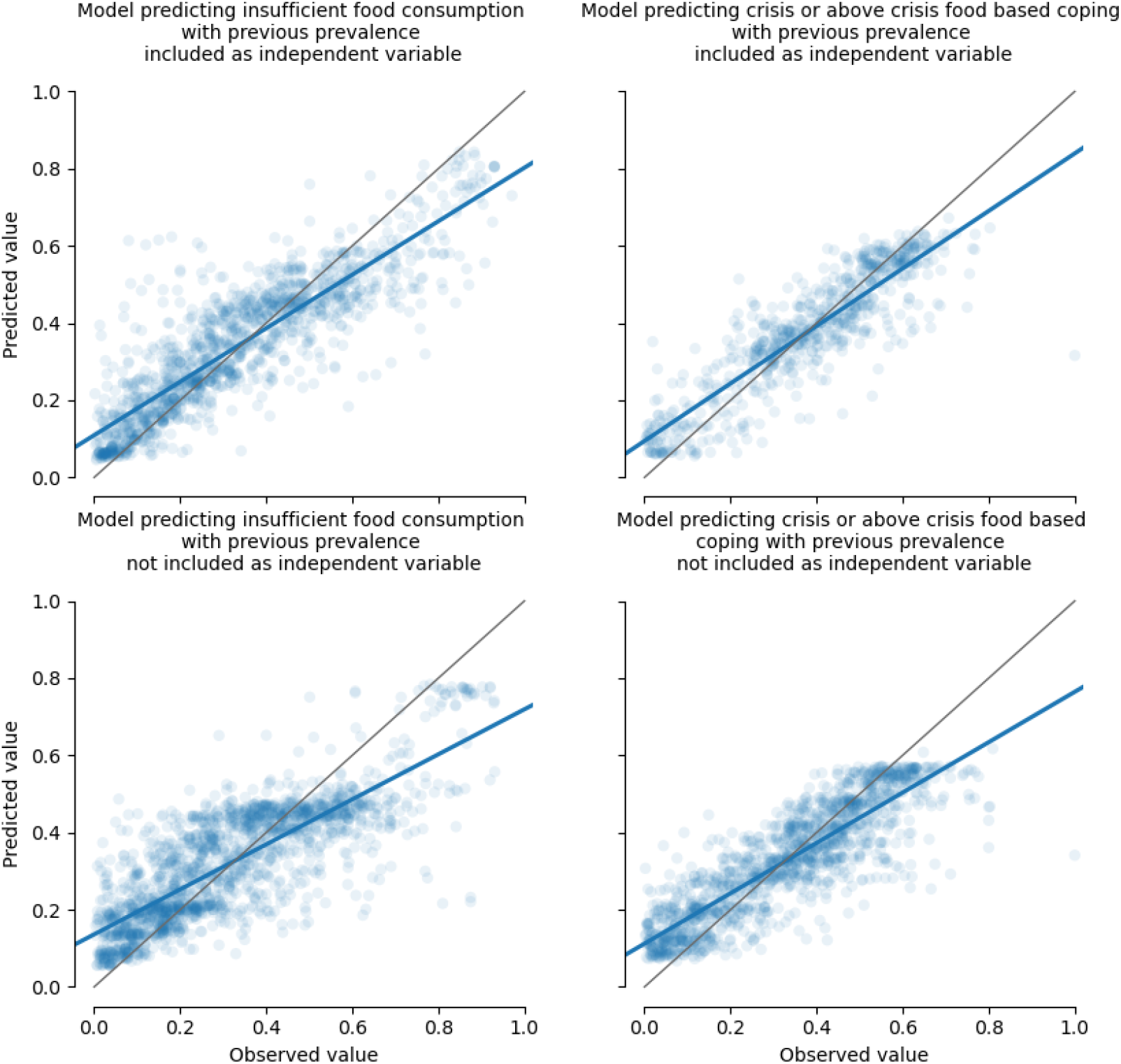
Predicted vs observed values. Each plot shows the predicted value (obtained as the median of *N*_*b*_ = 100 predictions, each generated from one of the bootstrapped models) vs the actual value for each observation in the test set. The blue line represents the best fit for the plotted points, whereas the gray line represents where the points would fall if all predicted values perfectly matched the observed ones. The closer the two lines are, the better the model’s performance is. The top plots refer to the models that include the prevalence from the previous assessment as an independent variable, and the bottom plots refer to the models that use secondary data only.

As one might expect, in both models the most predictive variable is the previously measured prevalence (see Supplementary Figure 1). Therefore, the question arises whether the independent variables built from secondary data bring significant additional information into the models or whether most of their explanatory power could be due to the previous assessment variable. To tackle this question, we compare the results of the proposed models with those obtained from a naive approach that simply uses the prevalence measured during the previous assessment as the predicted prevalence. We find that this naive model can explain only 39% of the variation in the prevalence of people with insufficient food consumption and 42% of the variation in the prevalence of people using crisis or above crisis food-based coping, demonstrating the fundamental importance played by secondary data to capture the dynamic nature of food insecurity and to explain the current situation when up-to-date primary data is not available.

### Predicting levels of insufficient food consumption and of crisis or above food-based coping from secondary data only

Having demonstrated the potential of the proposed approach when information on both key drivers and previous values of the target indicator is available, as well as the fundamental role played by the secondary data, we then tested to what extent insufficient food consumption and crisis or above food-based coping levels can be predicted when previously measured prevalence is not available.

We trained two additional models, using the same approach but removing the prevalence from the previous assessment from the set of independent variables, hence using secondary data only. As reported in Table 1, in this case results show that the proposed models are able to explain, on the test set, 63% of the variance in the prevalence of people with insufficient food consumption and 73% of the variance in the prevalence of people using crisis or above crisis food-based coping, with a mean absolute error of 0.09 and 0.07, respectively. Figure 2 (bottom plots) shows the predicted vs actual prevalence for each observation in the test set. As expected, these latter models have lower explanatory power and slightly higher errors than the former ones, however the reported metrics are still satisfactory. The advantage of these models is that they allow to predict the food security situation also in areas where no previous measurement is available, substantially expanding the application horizon of the proposed approach.

### Nowcasting levels of insufficient food consumption and of crisis or above food-based coping in near real-time

WFP is currently monitoring the food security situation in near-real time in a number of countries, collecting food consumption and food-based coping data through daily remote phone surveys [1, 22]. The predictive models proposed in this study aim at serving WFP’s need to estimate the situation in additional countries where primary near real-time data is not currently available, in order to provide humanitarian stakeholders with regular and frequent up-to-date global overviews of the food security situation and allow for timely decision making on resource allocation.

In order to test the effectiveness of the proposed models in capturing the current situation, we compared insufficient food consumption and crisis or above food-based coping trends measured by WFP’s near real-time monitoring systems between March 1^st^ and April 30^th^, 2021 with the corresponding prevalence predicted by the proposed models, which were trained and tested on data collected before March 1^st^, 2021. For areas where the prevalence from a previous assessment - performed prior to the start of the near real-time monitoring system in the country - is available, we use the models that include this information as an independent variable; for areas where this is not available, we resort to the models that rely on secondary data only.

National-level results for insufficient food consumption are shown in Figure 3. The red lines represent the target prevalence as measured by WFP’s near real-time monitoring systems, the blue lines the predicted prevalence, and the green dashed lines the prevalence from the previous assessments, where available. All prevalence levels were first obtained at the spatial resolution of first-level administrative units and then aggregated to obtain national trends. Sub-national trends are reported in Supplementary Figures 2-16. We can observe that in most cases the prevalence measured by the near real-time monitoring systems falls within the prediction intervals (or within a reasonable distance of less than 5%) for at least part of the trend. In those cases where the actual data line is further from the prediction interval, we can observe that the predicted trend is however significantly closer to the observed one than the prevalence from the previous assessment (e.g. Benin and Malawi). In the remaining case, where no previous assessment is available (i.e. Syria), the predicted and observed trends both fall within the same severity level (*>* 40%) defined by WFP [22]. Similar results can be observed for crisis or above food-based coping in Figure 4 (see Supplementary Figures 17-31 for the corresponding sub-national trends), with the exception of Niger.

**Figure 3:**
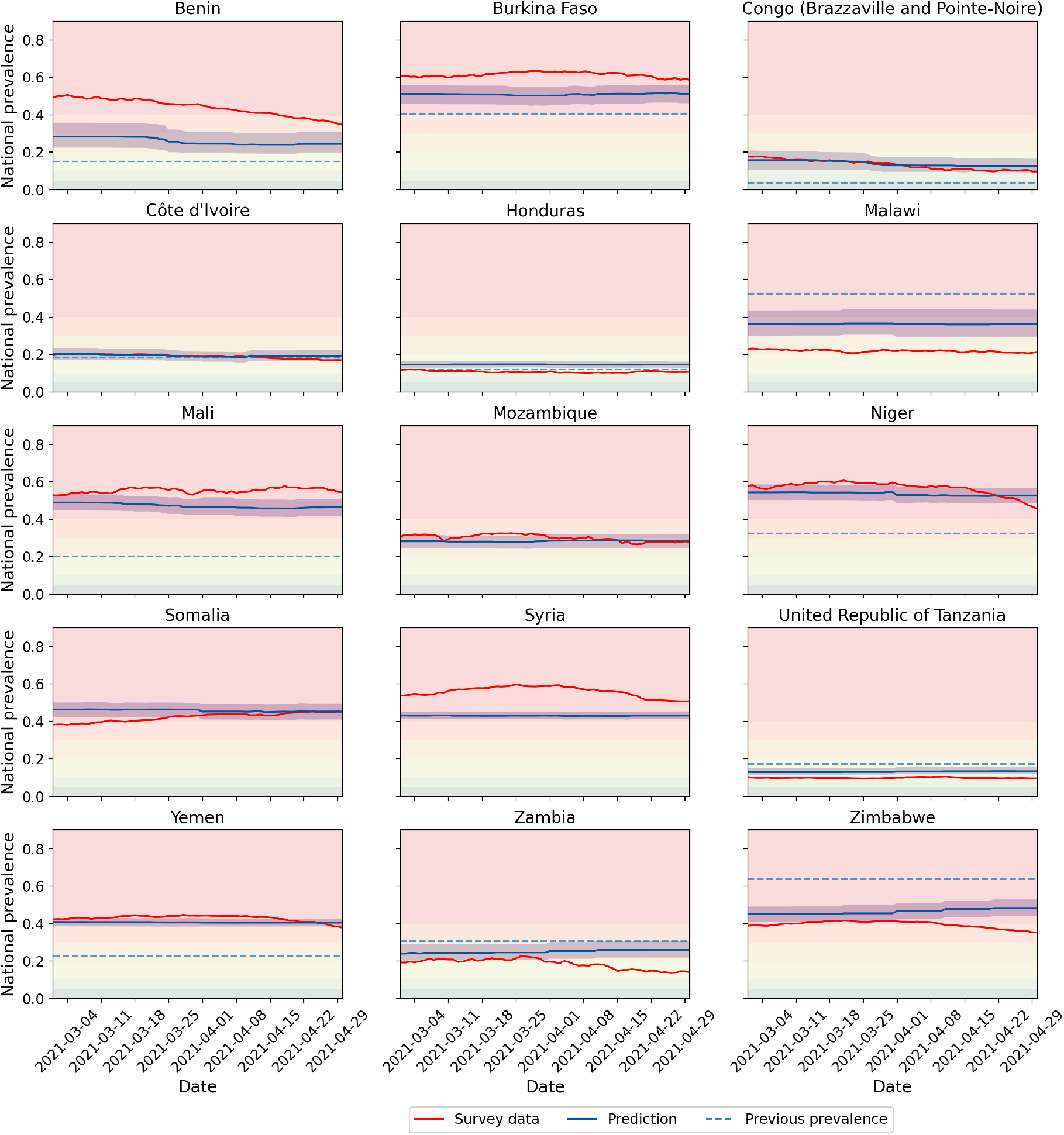
Comparison between near real-time monitoring of insufficient food consumption and predicted trends. Each plot shows the prevalence of people with insufficient food consumption between March 1^st^ and April 30^th^ 2021, as measured through WFP’s near real-time monitoring systems (red lines) and as predicted by the proposed models (the blue lines represent the median of the *N*_*b*_ = 100 bootstrapped models predictions and the light blue area around them the corresponding 95% confidence interval). The dashed green lines represent the value measured during the previous assessments (performed prior to the start of the near real-time monitoring system in the country), where available. The background colors represent severity levels as defined by WFP (*<* 5%: very low, 5 − 10%: low, 10 − 20%: moderately low, 20 − 30%: moderately high, 30 − 40%: high, *>* 40%: very high) [22].

**Figure 4:**
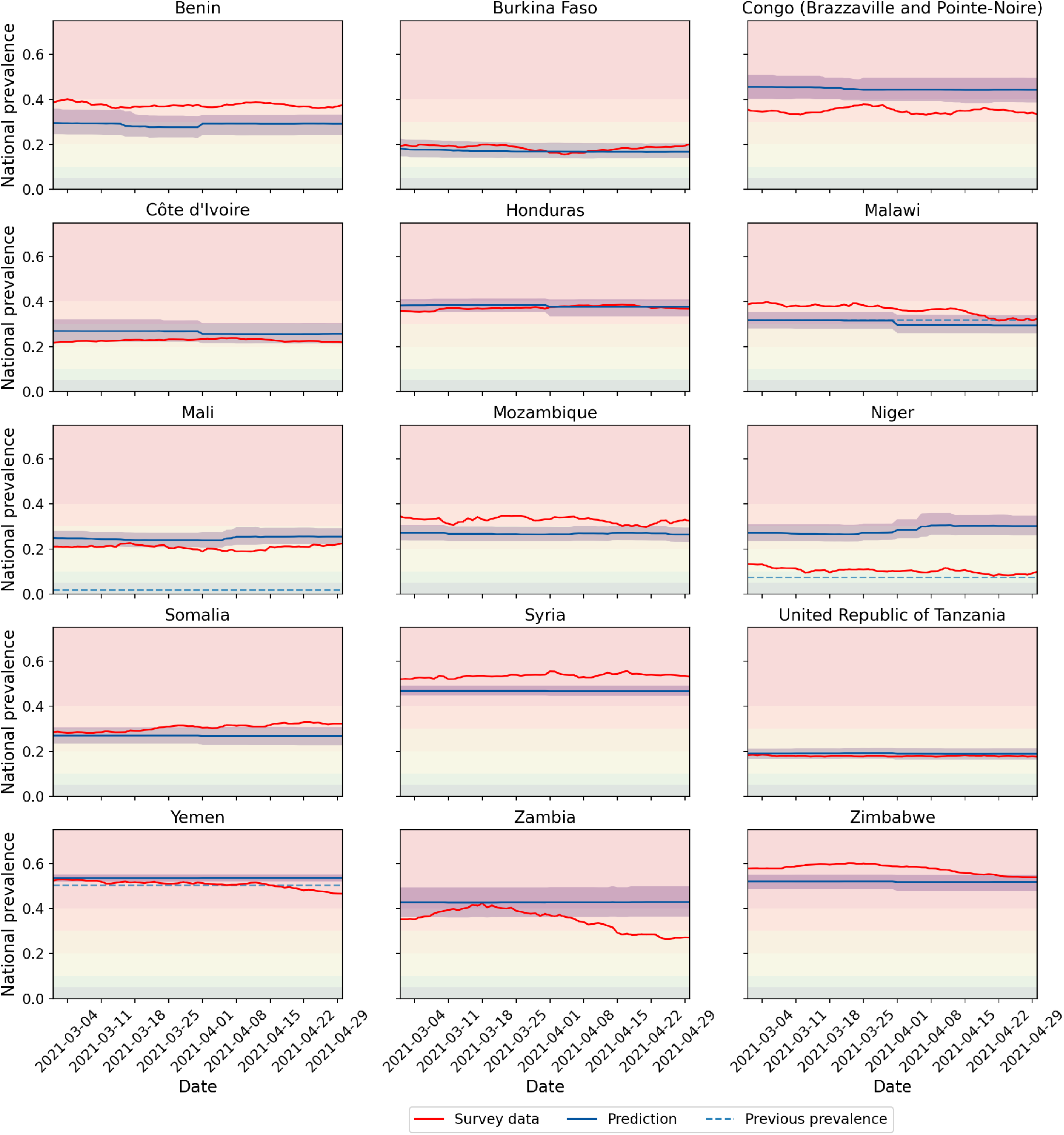
Comparison between near real-time monitoring of crisis or above food-based coping and predicted trends. Each plot shows the prevalence of people using crisis or above crisis food-based coping between March 1^st^ and April 30^th^ 2021, as measured through WFP’s near real-time monitoring systems (red lines) and as predicted by the proposed models (the blue lines represent the median of the *N*_*b*_ = 100 bootstrapped models predictions and the light blue area around them the corresponding 95% confidence interval). The dashed green lines represent the value measured during the previous assessments (performed prior to the start of the near real-time monitoring system in the country), where available. The background colors represent severity levels as defined by WFP (*<* 5%: very low, 5 − 10%: low, 10 − 20%: moderately low, 20 − 30%: moderately high, 30 − 40%: high, *>* 40%: very high) [22].

### Explaining predicted values and changes in predicted trends

Machine learning approaches are often seen as black boxes that provide recommendations without the user being able to access the process and rationale that generated them. This is not an acceptable practice when the model outputs are being generated in support of decision making. Therefore, in this context, proposing methods to explain predicted results is as important as building the models themselves.

Here, we make use of SHAP values [24, 25] to explain how each prediction is obtained. SHAP values make it possible to explain each predicted prevalence as a value obtained starting from the average prevalence observed in the training set (baseline) and then accounting for how much each independent variable contributes to the final prediction by moving the prevalence towards lower or higher values.

We first show how this method is able to demystify the predicted prevalence of people with insufficient food consumption or of people using crisis or above crisis food-based coping, explaining how the models predict values compatible with what has been measured through WFP’s near real-time monitoring systems. In Figure 5 we show an exemplification of this approach to explain both predicted indicators in the cases of Mali on March 1^st^, 2021, using a waterfall plot approach [25]. Starting from the bottom, each variable’s contribution is summed to the baseline *E*[*f* (*x*)] to eventually reach the predicted value *f* (*x*). Variables are ordered by importance (in terms of the absolute value of their contribution), and colored by the sign of the contribution: red if increasing and blue if decreasing, with respect to the baseline. In panel (a) we see that the most important variables in determining the high prevalence of people with insufficient food consumption (0.48) in Mali is the prevalence of undernourishment (*>* 5%), together with the low GDP per capita (793.5 USD). Conversely, the lower value of the prevalence measured during a previous assessment (0.20) drives the predicted value down, similarly to what happens in panel (b) when considering the prevalence of people using crisis or above crisis food-based coping.

**Figure 5:**
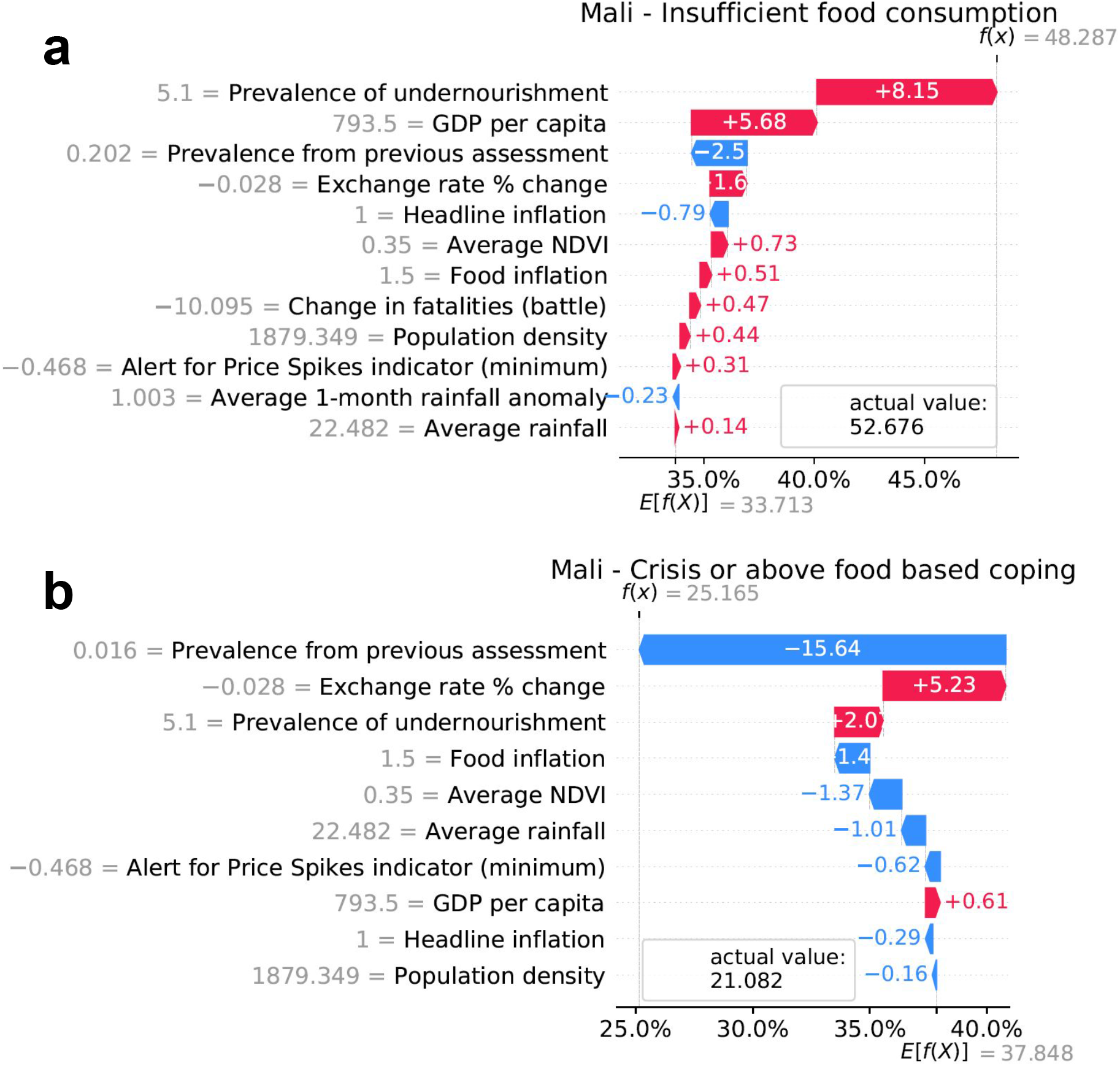
Explaining a single prediction. Waterfall plots [25] show how SHAP values are used to explain the predicted prevalence *f* (*x*) on March 1^st^ 2021 in Mali (aggregating first-level administrative unit predictions) as the sum of a baseline *E*[*f* (*x*)] and each variable’s contribution, highlighting in red positive contributions and in blue negative ones. Prevalence and variable contributions are expressed as percentages. Next to each variable’s name, its value averaged weighting by population over all first-level administrative units in the country is shown. The boxes contains the actual values measured through the near real-time monitoring system for the same day. Panel (a) shows the prevalence of people with insufficient food consumption and panel (b) of people using crisis or above crisis food-based coping. The corresponding sub-national plots are reported in Supplementary Figures 32-49.

Beyond using SHAP values to explain individual predictions, in this study we propose a novel method based on this framework to measure the relative importance of each independent variable in explaining predicted changes in food consumption and food-based coping between two dates. This is important for decision makers to understand why the models, when deployed to produce near-real time trends, show improvements or deterioration in the food security situation. This is done by exploiting the differences in SHAP values between two dates and its mathematical formulation is detailed in the *Methods*. In Figure 6 we show the proposed method applied to two specific examples. On the left, we see how the predicted food consumption situation in Lesotho slightly deteriorated between March 1^st^ and April 30^th^, 2021. Our method is able to identify that the most important variable in determining this change has been the anomaly in precipitation with respect to the historical average in the same period. As shown in the bottom table, rainfall anomaly went from 111% (above average, i.e. it rained more than usual) to 80% (below average, i.e. it rained less) within the two month period under consideration. Other variables also had a smaller impact in the change, for example an increase in both food and headline inflation also caused a deterioration, while a decrease in the price alert indicator (which measures changes in cereal prices) and a stabilization of the currency exchange rate both caused a small improvement in the situation. Let us note that variables which did not change their value during the time period considered can however still change their SHAP importance, as this is relative to the values of all variables at each point in time. On the right, we explain a deterioration in the predicted prevalence of people using crisis of above crisis food-based coping in the same time period in South Sudan. In this case, the main causes are a decrease in the average rainfall and an increase in the price alert indicator. Sub-national investigations reveal that the former is the main driver in areas like Western Bahr el Ghazal or Western Equatoria, whereas the latter is responsible for changes in other areas like Lakes and Jonglei, as one can see in Supplementary Figures 60-69.

**Figure 6:**
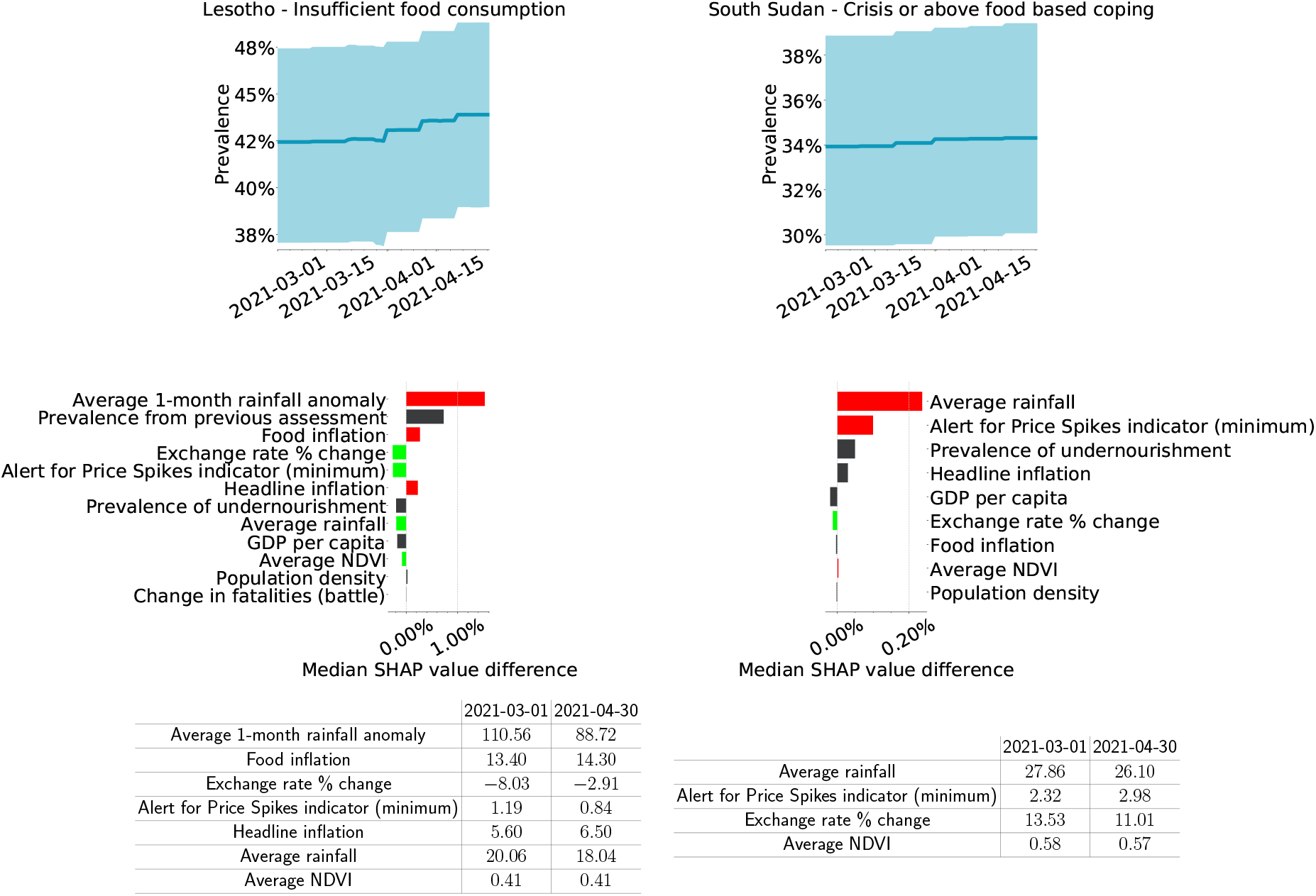
Explaining changes in predicted trends between two dates. A SHAP values based method was developed to explain why the models are predicting changes in prevalence between two dates. On the left we show the predicted prevalence of people with insufficient food consumption in Lesotho between March 1^st^ and April 30^th^, 2021. On the right, the predicted prevalence of people using crisis or above crisis food-based coping in South Sudan in the same time period. On the top plots we can see the predicted trends in blue, with the light blue area around them the corresponding 95% confidence interval. The middle plots show each independent variable’s contribution to the change: positive contributions (deterioration) are shown in red, negative contributions (improvements) in green, and variables which did not change value between the two dates are shown in gray. Variables are ordered by importance (in terms of absolute value of their contribution). All prevalence and SHAP value differences are expressed as percentages. The tables in the bottom report the values of the models’ independent variables at the two considered dates. The corresponding sub-national plots are reported in Supplementary Figures 50-69.

## Discussion

In this study, we propose an approach that makes it possible, for the first time, to predict the current sub-national food consumption and food-based coping situation on a global scale from secondary data on the key drivers of food insecurity. We show that when a previous measurement of the target indicator is available and included as independent variable, the models, as expected, have a higher explanatory power and lower errors than when relying on secondary information only. Importantly, we also demonstrate that these models perform significantly better than a naive approach that uses the prevalence measured during a previous assessment as the predicted prevalence. Moreover, even the models that rely on secondary information only show a satisfactory explanatory power. Having trained and validated the proposed models on historical data, we then further show that they can be used to predict the current prevalence of insufficient food consumption and of crisis or above food-based coping, by comparing the data being collected in near real-time by WFP during a recent two month period with the corresponding predicted levels. Finally, we show that, despite the non-linear tree-based model structure, it is possible to provide interpretable explanations of predicted figures and of what causes changes over time, even if the models do not have an intrinsic dynamic component.

Despite the encouraging results, several limitations apply. First of all, the proposed models are trained and validated combining together sub-national data from a number of different countries. Having such rich variety of data made it possible to build a global model that can be used to estimate the food security situation in any area in the world. However, this also means that the models learnt patterns in the data that correspond to what is most commonly observed across the different countries, limiting the discovery of less frequent patterns specific to local contexts. These latter patterns would be more easily discovered by training separate models each based on historical data from a specific country only. This would however require the availability of large enough samples for each individual country, which is currently not available for a number of countries. Hence the reason for the proposed approach. However, it should be noted that predictions generated by the proposed models should be used with caution and further validated when relative to areas and countries that are not represented in the historical data used to train and test the models, as discussed in previous studies [26]. In this sense, one of the challenges faced in the model development was the unequal availability of food consumption and food-based coping data across different countries. In order to ensure as much as possible a balanced geographical representation, we resorted to sampling and only kept a subset of the available data for the most data-rich countries, while also ensuring enough data was included to properly train the models.

In regard to the secondary information feeding the proposed models, in this study we resort to data on the three main drivers of food insecurity. Undoubtedly, further information could be included in order to enrich the models, such as data on displacements, natural hazards, animal and crop diseases and epidemic outbreaks [10]. However, the limited availability of relevant data on a global scale and on a multi-annual time frame restricts the possibilities of expansion to additional independent variables. Given that the time frame covered by the historical data used to train and test the models includes the COVID-19 pandemic, one might expect that we would need to include this information in the models, for example in the form of caseload or death incidence. This is however not the approach we adopted, since the objective was to build a general model for food insecurity, not specific to the current situation. Our assumption, which is confirmed by the high explanatory power obtained even without taking COVID-19 explicitly into account, is that the effects of this pandemic on the food security situation are already indirectly taken into account by some of the independent variables included in the models, namely those accounting for staple food prices in local markets and macro-economic indicators. Further investigation should however be the subject of future work.

Some challenges and limitations also apply to the secondary data that has been incorporated in the models to build the independent variables. First, the different data sources do not have the same time resolution and update frequency, which range from annual estimates to daily measures, as further detailed in the *Methods*. This means that, when generating predictions on a regular basis to nowcast the situation, most variables will not update daily and therefore in order to see significant changes longer time intervals need to be considered. Secondly, spatial resolution also varies across the different data sources. Whereas population density, rainfall, vegetation and conflict-related fatalities data are available at the first level administrative unit resolution, macro-economic indicators and prevalence of under-nourishment are national figures, leading to all sub-national areas being assigned the same value. This could be seen as a limitation but it also allows to provide to each first-level administrative unit some national characterization, which would otherwise be lost in a global model trained with sub-national level data only. Let us consider, for example, two bordering areas belonging to two different countries, such as Venezuela and Brazil. Given their geographical proximity, they might be highly similar with respect to vegetation and precipitations, but, concurrently, they would be highly different in terms of the economic situation, possibly resulting in profound differences in the food security situation.

Having carefully taken into account the challenges and limitations highlighted above, the proposed models have the potential to be used to provide unique information to humanitarian decision makers when no primary data is available. Predictions should certainly be handled with caution and never considered as ultimate truth. When indicating some level of deterioration, they should serve as triggers for rapid assessments and more in-depth analysis of the situation, rather than being used to prompt immediate decision making. In this regard, the proposed methods give decision makers more insights into how the model predicted a certain figure or changes in the predicted trends, allowing for a deeper understanding of the situation. Finally, it should be noted that, in order to ensure continued validity of the proposed models, it is essential to perform regular re-trainings whenever a significant amount of new primary data is collected and available. This will allow to improve the models explanatory power thanks to the increased volume and variety of data the training is performed on, as well as to eventually learn new emerging patterns, hence remaining representative of the current situation.

## Methods

### Target indicators

The two target indicators of the proposed predictive models - the prevalence of people with insufficient food consumption and the prevalence of people using crisis or above crisis food-based coping - are calculated on the basis of two household-level indicators: the Food Consumption Score (FCS) [27] and the reduced Coping Strategy Index (rCSI) [28], respectively.

The FCS is calculated by asking each household how often, during the last 7 days, they have consumed items from the different food groups: main staples, pulses, vegetables, fruit, meat and fish, milk, sugar, oil and condiments. Each consumption frequency is then weighted according to its relative nutritional importance to obtain the *FCS* = ∑*w*_*i*_*x*_*i*_, where *w*_*i*_ is the weight of food group *i* and *x*_*i*_ the frequency of its consumption by the household, that is the number of days for which the food group was consumed during the past 7 days. Once the food consumption score is calculated, each household is then assigned a food consumption group (poor, borderline or acceptable) based on standard thresholds, which can however be adapted based on specific consumption behaviours in the country of interest. Food group weights and thresholds are detailed in [27]. The prevalence of people with insufficient food consumption is then obtained as the prevalence of households in the sample with poor or borderline food consumption [29].

To compute the rCSI, households are asked if and how often during the last 7 days they had to adopt the following coping behaviors: relying on less preferred or less expensive food, borrowing food from relatives or friends, limiting portion sizes, restricting adults’ consumption in order for small children to eat and reducing the number of meals eaten in a day. Coping strategy frequencies are then weighted according to their severity to obtain the rCSI, as detailed in [28]. The prevalence of people using crisis or above crisis food-based coping is then obtained as the prevalence of households in the sample with rCSI ≥ 19 [29].

The available historical data for the two indicators is at the spatial resolution of first-level administrative units and has been collected through both face-to-face and mobile phone surveys, including those from WFP’s near real-time monitoring systems. The insufficient food consumption data spans units across 78 countries from 2006 to February 2021, and the crisis or above food-based coping data units across 41 countries from 2013 to February 2021, with more than 200,000 observations for each indicator. This large volume of data is however not equally representative of all covered geographical areas: countries where a WFP’s near real-time monitoring system is in place are over-represented since these systems provide data on a daily basis, whereas in the remaining countries data collection is performed only a few times per year. Therefore, in order to avoid training the models on an unbalanced data set, sampling is performed by randomly selecting, for each first-level administrative unit, one observation per month only. The final data set used to train and test the models where the prevalence from previous assessment is not included as independent variable is composed of 8737 observations for food consumption and 7031 for food-based coping. For the models where the prevalence from a previous assessment is instead included, the size is further reduced because only observations preceded by a previous one can be used, resulting in 6619 observations for food consumption and 3499 for food-based coping. The breakdown by country of all of the above mentioned numbers is reported in Supplementary Table 1.

### Modeling approach

The statistical approach adopted in this study is logistic regression, as the proposed models predict the probability of having a person with insufficient food consumption or using crisis or above crisis food-based coping in a given area at a given time. Gradient Boosted Decision Trees [30] were identified as the most suitable algorithm to perform the regressions, given its high performance, flexibility, and its capacity to handle complex and non-linear relationships. The XGboost implementation was used [23].

Four different models were developed: two for the prevalence of people with insufficient food consumption and two for the prevalence of people using crisis or above crisis food-based coping. In each case, one includes the prevalence from a previous assessment as independent variable and one does not.

For each model, a 20% random sample of the data was used as test set, while the remaining 80% was used for tuning the hyper-parameters via a 4-folds cross-validation, and for training the models. The tuned hyper-parameters and the explored values are listed in Supplementary Table 2. The chosen combination of hyper-parameters is the one leading to the smallest difference between the average *R*^2^ on the folds used as training set and the average *R*^2^ on the folds used as validation. We opted for this criterion to favor models where the performance on the test is the most similar to the performance on the training set, since large differences are often an indication of overfitting. Once the hyper-parameters are selected, *N*_*b*_ = 100 models are fitted on samples with replacement of the training set (i.e. bootstrapping) and the test set is used to evaluate the model’s performance. For each observation in the test set, *N*_*b*_ predictions are generated (one per bootstrapped model), and the median value is then used to calculate the model performance metrics, i.e. the coefficient of determination *R*^2^ and the mean absolute error (MAE). Supplementary Figures 70-73 show that convergence for both metrics is reached within 100 bootstraps.

### Feature definition

The initial set of considered features is composed of variables related to food insecurity and its main drivers: economic shocks, extreme weather events and conflict [10].

#### Food prices

To capture variations in cereal and tubers prices, we resort to the Alert for Price Spikes (ALPS) indicator [31]. This metric is based on a trend analysis of monthly price data: the idea is to compare the long-term seasonal trend of a commodity’s price time series in a market with the last observed price in the same market, providing an indication of the intensity of the difference between the current market price and the historical trend. The higher the difference, the more severe the alert. Price data and the corresponding ALPS calculations are publicly available through WFP’s Economic Explorer platform [32]. If more than one market is present within a geographical area, the average ALPS value is considered. If no market is instead monitored in a given area, the national average is considered. From this data, we build a set of three features by taking into account the minimum, maximum and average ALPS value within a three month window. The length of the window was selected as the shortest time period that minimizes the number of missing values in the training and test set. A one month lag is applied to ensure data availability when deploying the model in real-time.

#### Macro-economic indicators

The following four macro-economic features are considered: most recent available annual GDP per capita in a four years time window, most recent available monthly headline and food inflation rates in a six months time window (applying a one month lag), and percentage variation between the average value of the currency exchange rate during the last three months and the average value during the previous three, to capture main changes in the situation. The three month window was selected for consistency with the food price features and the same applies to the following features too. Being all country-level indicators, the same national value is assigned to all first level administrative units within a country. Data is obtained from Trading Economics, a website providing publicly available economic and financial indicators, including historical data, for 196 countries [33]. For countries where unofficial currency exchange rate is collected by WFP, these values are used instead of official ones [34], since they provide a more reliable representation of the country economic situation.

#### Weather

An initial set of five weather features is built by taking: i) the average rainfall and normalized difference vegetation index (NDVI) during a 12 month time window, which allow to characterize each first-level administrative unit’s climate; ii) rainfall and NDVI anomalies with respect to historical averages. For rainfall, two anomalies have been defined by WFP: the ratio between the amount of rainfall during the last one month or three months and the historical average of the amount of rainfall in the same period of the year. For NDVI, a single anomaly is defined based on the last 10 days, since vegetation already integrates the effects of previous rainfall. All three anomalies are provided for each 10-day window of the year, and we take their average during a three month window like for previous features, applying a 10-day lag. Data is obtained from WFP’s Seasonal Explorer platform [35], which provides open rainfall and NDVI time series for a near-global set of administrative units, computed, respectively, from CHIRPS [36] and MODIS [37] data.

#### Conflict

Conflict data is obtained from the Armed Conflict Location & Event Data Project (ACLED), a publicly available repository of reported conflict events and related fatalities across most areas of the world [38]. The date, latitude and longitude of each event is reported, allowing to match it to the corresponding first-level administrative unit. In order to capture deterioration or improvements in the conflict situation, we define the conflict features as the difference between the number of reported fatalities during the last three months and the three months prior, applying a 14-day lag. Only fatalities reported in events involving organized violence (i.e. “battles”, “violence against civilians”, and “explosions/ remote violence”) are considered [39], and a total of seven features is obtained by considering these three categories separately and in combination.

#### Prevalence of undernourishment

The most recent available prevalence of undernourishment in a 4 year time window is considered. This is a national yearly indicator publicly available in FAOSTAT [40]. Being a country-level indicator, the same national value is assigned to all first level administrative units within a country.

#### Population density

Yearly population density is also considered and obtained from CIESIN raster files [41] by averaging all pixel values within each first-level administrative unit. Estimates for years not covered by the data set are obtained through linear interpolation.

#### Previous value of the target indicator

Finally, the previously measured prevalence of people with insufficient food consumption and of people using crisis or above crisis food-based coping are also considered, when available. For first-level administrative units where WFP’s near real-time monitoring is in place, only data collected prior to the start of the near real-time monitoring is used to build this feature. This choice was made because the proposed models are meant to be used in practice in situations where no near real-time monitoring is in place, and hence the last available value would be from a face-to-face or mobile phone assessment conducted in a specific and limited time window in the past.

### Feature selection

Having defined an initial set of features, a selection process was then performed to single out the final sets of independent variables that maximize models’ performance. First, we carried out a backward feature elimination process for each of the 4 models. We first trained them with the initial set of features, then removed the one with the lowest importance. Feature importance is defined as the gain across all the tree splits in which a certain feature is used [23]. At each step we then re-performed the training and eliminated the least important feature. This process is repeated *N*_*i*_ = 500 times, each time using different 80%/20% random samples to train and test the model. The number of iterations was chosen to ensure the convergence of model performance metrics on the test set, as shown in Supplementary Figures 74-77. This first elimination process results in a ranking of the features from most to least relevant, by summing up, for each, its model importance values across all steps and iterations. Following this ranking, a forward selection process is then performed. The model is first trained with the most important feature only and then, at each step, one feature is added following the ranking, and the model is re-trained. Each step is performed *N*_*i*_ = 500 times using different random samples to train and test the model. Finally, for each step the average values of the model performance metrics on the test set across the *N*_*i*_ iterations are computed and the final set of features is chosen as the one maximizing the *R*^2^ and minimizing the MAE (see Supplementary Figures 78-81). The final list of features used as independent variables in each model are reported in Supplementary Table 3.

### SHAP values

SHAP (SHapley Additive exPlanations) [24] is a framework recently proposed to interpret predictions made by often complex black box machine learning algorithms. SHAP unifies other methods (Lime, DeepLift, etc.), and for tree-based models it allows to write a prediction as the sum of a baseline value and each feature’s contribution [25]:

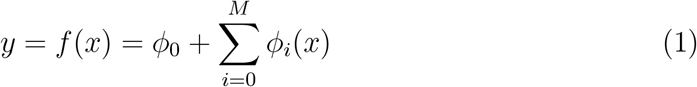

SHAP values for tree-based models like XGboost have been shown to improve on other local tree explanations, like visualizing the decision tree, not feasible for tree ensembles, or model-agnostic local explanations, computationally expensive if explaining large data sets [25]. Moreover, SHAP local explanations can be used as building blocks for global explanations, as shown in Supplementary Figure 1, where we take the mean absolute value of SHAP values across all data points to build a global feature importance ranking. In this study, we use the python open-source implementation of the TreeSHAP algorithm [42].

### Explaining the single prediction

SHAP values represent each feature’s contribution towards the model prediction, and their absolute value can therefore be interpreted as each feature’s importance. This method improves on widely used global feature importance methods like split-based or gain-based measures, as it allows to compute prediction-specific feature importance. As detailed in previous sections, each of our four models actually consists of *N*_*b*_ = 100 different models fitted on different samples (with replacement) of the training data, of which we report the median prediction and confidence interval. To determine the importance of a feature we then take each feature’s median SHAP value across the *N*_*b*_ bootstrapped models. Convergence checks are reported in Supplementary Figures 82-83.

In this study, predictions are originally obtained at the spatial resolution of first-level administrative units, but they can then be aggregated to display results at the national level too. To determine features importance at the country level, we average the SHAP values across all first-level administrative units in a country, by weighting each value according to the unit’s population. This can be easily interpreted: a SHAP value corresponds to the change in prevalence, with respect to the baseline, due to one feature. By performing a population-based weighted average we are computing the change in number of people due to that feature. This operation then sums the number of people across all units, and divides it by the country population to return the change in national prevalence. The same operation is also performed on the model baseline. Note that this also allows us to combine predictions coming from areas with and without a previous value of the target variable, even if the underlying models use slightly different sets of features.

### Explaining trend changes

This method allows us to compute the feature importance ranking for a given area in a given day, explaining which features were the most important and how they contributed to the final prediction. However, predictions for the same area can produce a trend in time, which in turn can show changes in prevalence due to changes in the input variables. We extend the SHAP framework to explain which features are responsible in determining changes in predicted trends.

Let us take two predictions *y*^*t*1^ and *y*^*t*2^ corresponding to the same area but two different dates. Following Eq. 1, we can write the trend change *y*^*t*2^ − *y*^*t*1^ in terms of the change *χ*_*i*_ in SHAP values relative to each of the *M* features:

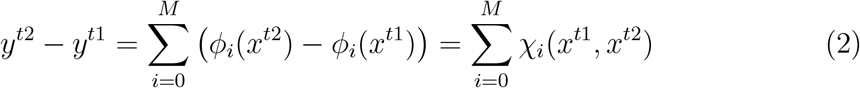

The features with largest associated SHAP value change are the ones that determined the trend change. Moreover, the sign of the change *χ*_*i*_ also tells us whether that feature has caused an increase or decrease in the prevalence, that is a deterioration or improvement in the food security situation.

Note that Eq. 2 is exact when considering a single model for a single first-level administrative area, but is only an approximation when considering median SHAP values, as previously mentioned. It is also important to note that this method can give apparently misleading indications due to nonlinear interactions between features. For instance, a feature that does not change value between the two dates can be the one with the largest SHAP value change (i.e. determining the trend change). This happens because other features change value, thus changing its relative importance in the two predictions. One could overcome this limitation by computing SHAP interaction values [25], but the computation is not feasible when dealing with our sample size, i.e. 400 models (100 bootstrapped iterations per model) and daily computations. Moreover, this would imply dealing with an order of 50 (number of features squared, divided by 2) different interactions, which would greatly undermine our effort to produce explainable predictions for decision makers.

## Data Availability

An API for insufficient food consumption and crisis or above food-based coping data is being developed by WFP. All other data is already publicly available and sources are referenced in the paper.

## Supplementary Information

Supplementary figures and tables are available here.

## Disclaimer

The content and views expressed in this paper are solely those of the authors and do not necessarily reflect the official views of the UN World Food Programme.

## Acknowledgements

The authors would like to thank their colleagues at WFP in HQ, Regional Bureaus and Country Offices for the fruitful discussions, and Alibaba Cloud for their initial contribution in terms of data infrastructure and modeling approach.

## Author contributions

EO, LR and JR conceptualized the study, GM and AB contributed to its further design. GM, LR and EO analyzed the data. GM, AB, SJ, MC, LR and EO designed and developed the python code used in the work. EO supervised the research. EO, GM and AB wrote the manuscript, all authors reviewed and approved it.

